# Rapid and Quantitative Detection of Human Antibodies Against the 2019 Novel Coronavirus SARS CoV2 and its Variants as a Result of Vaccination and Infection

**DOI:** 10.1101/2021.07.13.21260442

**Authors:** Benjamin Taubner, Ruben Peredo-Wende, Ananthakrishnan Ramani, Gurpreet Singh, Klemen Strle, Nathaniel C. Cady

**Author notes:** Corresponding Author: Nathaniel C. Cady, PhD, SUNY Polytechnic Institute.

## Abstract

Measuring the antibody response to 2019 SARS CoV2 is critical for diagnostic purposes, monitoring the prevalence of infection, and for gauging the efficacy of the worldwide vaccination effort COVID-19. In this study, a microchip-based grating coupled fluorescent plasmonic (GC-FP) assay was used to measure antibody levels that resulted from COVID-19 infection and vaccination. In addition, we measured the relative antibody binding towards antigens from variants CoV2 virus variants, strains B.1.1.7 (UK) and B.1.351 (S. African). Antibody levels against multiple antigens within the SARS CoV2 spike protein were significantly elevated for both vaccinated and infected individuals, while those against the nucleocapsid (N) protein were only elevated for infected individuals. GC-FP was effective for monitoring the IgG-based serological response to vaccination throughout the vaccination sequence, and could also resolve acute (within hours) increases in antibody levels. A significant decrease in antibody binding to antigens from the B.1.351 variant, but not B.1.1.7, was observed for all vaccinated subjects when measured by GC-FP as compared to the 2019 SARS CoV2 antigens. These results were corroborated by competitive ELISA assay. Collectively, the findings suggest that GC-FP is a viable, rapid, and accurate method for measuring both overall antibody levels to CoV2 and relative antibody binding to viral variants during infection or vaccination.

## INTRODUCTION

The 2019 novel coronavirus, SARS CoV2 (COVID-19) has resulted in millions of deaths worldwide, and has spurred the development of novel diagnostic strategies, detection technologies, and vaccination approaches. Monitoring an individual’s antibody responses to CoV2 antigens has become paramount from an epidemiological perspective, but also as a means of determining the efficacy of vaccination. By measuring the levels of antibodies (primarily IgG) in human blood or other bodily fluids, an individual’s prior infection history, as well as their serological response to vaccination can be elucidated. Monitoring the stability and/or decline of antibody levels over time is important in estimating how long individuals will retain immunity (1).

Beyond assessing human serological response to infection or vaccination, it is important to understand how an individual’s immune system will respond to a growing number of novel CoV2 variants. A core area of concern is mutations in the spike protein, which could interfere with antibody binding, and subsequently affect the blockade (neutralization) of viral entry into human cells via the ACE2 receptor (2). This could ultimately result in breakthrough infections for individuals who were previously infected or vaccinated (2). Recent studies have shown that emerging variants in the United Kingdom (B.1.1.7) and South Africa (B.1.351) are not as effectively neutralized by blood serum from vaccinated individuals, nor from those who were previously infected with the original 2019 SARS CoV2 strain (2-4). This is also an area of concern for variants emerging in other regions, including Brazil and India (5, 6). This highlights the need for sensitive, specific, high-throughput assays to monitor antibody levels and predict effectiveness.

Multiplexed quantitative serological assays; allow both the determination of antibody levels in response to infection and vaccination, and the ability to assess relative antibody neutralizing capacity, provide an attractive diagnostic solution. Established methods of assessing antibody response to SARS CoV2 and its variants include cell-based viral neutralization assays (2, 3, 7, 8) and competitive *in vitro* binding assays such as ELISA (9, 10). Previously we demonstrated a grating-coupled fluorescent plasmonic (GC-FP) biosensor platform for rapid (30 min), quantitative, multiplexed detection of human antibody response to both Lyme disease and COVID-19 infection (11, 12). The GC-FP detection ratio (ratio of antibody binding to target antigens vs. negative control proteins) for human serum and dried blood spot samples correlated strongly with standard antibody testing approaches, including microsphere immunoassay (MIA) and ELISA (12). Notably, we found that dried blood spot samples as well as more traditional blood serum samples could yield high sensitivity and selectivity for diagnosing prior COVID-19 infection.

In the study presented here, we modified GC-FP detection microchips to include additional antigens from variant strains of SARS CoV2, including B.1.1.7 and B.1.351. Using these new detection microchips, we were able to simultaneously monitor antibody levels against multiple 2019 SARS CoV2 antigens for individuals throughout the vaccination sequence, and for all three vaccine types currently approved for use in the United States: Pfizer-BioNTech (13), Moderna (14) and Johnson & Johnson (15). We also assessed the relative binding of antibodies to original and variant strains of SARS CoV2 antigens for multiple exposure scenarios including: 1) acute severe (hospitalized) infection, 2) mild infections that did not lead to hospitalization, 3) vaccination with all three vaccines, 4) and a combination of prior infection and vaccination. Our results demonstrate that GC-FP is an effective and sensitive method to monitor antibody levels in response to vaccination, and can determine relative antibody binding levels to original and variant 2019 SARS CoV2 antigens, all in a single, rapid test.

## MATERIALS AND METHODS

### Materials

Nucleocapsid protein (N), the S1 fragment of the spike protein (S1), the extracellular domain of the spike protein (S1S2), the receptor binding domain of the spike protein (RBD) for the 2019 SARS CoV2 virus, S1 variant antigens, RBD variant antigens, and human serum albumin (HSA) were all obtained from Sino Biological, Inc. (Table 1). Positive control protein, human IgG protein (Hum IgG), SuperBlock blocking buffer and phosphate buffered saline (PBS) were obtained from ThermoFisher Scientific. PBS-TWEEN (PBS-T) solution consisting of PBS + 0.05% v/v TWEEN-20 (Sigma-Aldrich) was prepared on a daily basis for all experiments. Alexa Fluor 647 labeled anti-human IgG (heavy and light chain) were obtained from Invitrogen/ThermoFisher Scientific. ACE2 competitive ELISA testing was performed using COVID-19 ACE2 testing kits from RayBiotech (COVID-19 Spike Variant-ACE2 Binding Assay Kit).

**Table 1.** Proteins and peptides used for generating GC-FP detection microchips.

| Antigen/Protein | Source |
| --- | --- |
| Human IgG (positive control) | Thermo Fisher |
| Human Serum Albumin / HSA (negative control) | Sino Biological |
| 2019 SARS CoV2 Nucleocapsid (N) | Sino Biological |
| 2019 SARS CoV2 RBD (RBD) | Sino Biological |
| 2019 SARS CoV2 S1S2+ECD (S1S2) | Sino Biological |
| 2019 SARS CoV2 S1 (S1) | Sino Biological |
| SARS CoV2 RBD – B.1.1.7 (N501Y) | Sino Biological |
| SARS CoV2 RBD – B.1.351 (K417N, E484K, N501Y) | Sino Biological |
| SARS CoV2 S1 – B.1.351 (K417N, E484K, N501Y, D614G) | Sino Biological |
| SARS CoV2 S1 – B.1.1.7 (ΔHV69-70, ΔY144, N501Y, A570D, D614G, P681H) | Sino Biological |

### Grating-Coupled Fluorescent Plasmonic (GC-FP) Biosensor Chip Preparation

Gold coated grating-coupled fluorescent plasmonic (GC-FP) biosensor chips were fabricated as described our previous work (11, 12, 16). GC-FP chips were also printed as described previously (12). A map of the protein/antigen spots is shown in the supplementary data, Figure S1.

### Biological Samples

Serum samples were collected from COVID-19 patients admitted to Albany Medical Center between October and December 2020 who enrolled in a study “Defining genetic and immune factors in COVID-19 severity.” All patients had a positive RT-PCR test for CoV2 and were hospitalized due to severity of their illness. Sera were processed on the day of collection and placed into -80C freezer until analyses. The study was approved by the IRB at Albany Medical Center (Protocol # 5929).

Dried blood samples were collected by the finger stick method. Lancet devices (27 ga.) and Whatman 903 protein saver collection cards were sent to volunteers with instructions and consent form approved by the SUNY Polytechnic Institute Institutional Review Board (protocol #IRB-2020-10 and #IRB-2021-2). Blood droplets were collected, allowed to dry, and then either hand delivered or mailed (via US Postal Service) to SUNY Polytechnic Institute. Following receipt of DBS samples, a 6 mm diameter biopsy punch was used to remove samples from the collection cards, which were then soaked in 500 µl of PBS-T ∼12 hr at 4 °C with rocking. Samples were collected from 1) participants who had no known exposure to COVID-19 and/or tested negative for COVID-19 infection, 2) vaccinated individuals at different time points throughout vaccination sequence, including pre-vaccination, at the second dose, and two weeks after the second dose, 3) vaccinated individuals at a single time point, a minimum of 2 weeks after final vaccination dose, and 4) vaccinated individuals who were previously diagnosed with COVID-19 infection via RT-PCR. The age, gender, and exposure/vaccination information for all participants represented in this work are listed in supplementary tables S1 and S2.

### GC-FP Detection Assay and ACE2 Competitive Assay

GC-FP microchips were processed using the same conditions described previously (12). Total assay time from sample introduction to chip imaging was 30 min. For serum testing, a standard dilution of serum in PBS-T (1:50) was used. For dried blood spot testing, undiluted extract (from extraction in 500 µl PBS-T) from the 6 mm diameter segment of the blood collection card was used in place of serum. Ciencia image analysis LabView software was used to define a region of interest (ROI) for each individual spot on the GC-FP biosensor chip and the fluorescence intensity of each spot was measured. The fluorescence intensity of all spots was normalized to the human IgG (Hum IgG) internal control spots on each chip, to account for variability between individual chips and individual experiments, generating a “GC-FP detection ratio” for every protein/antigen included in the GC-FP microchip (12):

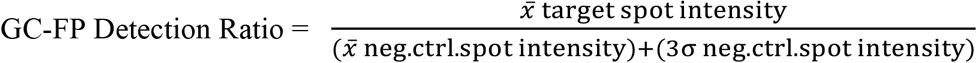

To determine if GC-FP antibody binding data for 2019 SARS CoV2 vs. B.1.1.7 and B.1.351 variant antigens was consistent with standard methods, an ELISA-based ACE2 competitive binding assay (Ray Biotech) was used, as per the manufacturer’s instructions. Additional details are provided in the supplementary information. Percent binding inhibition was calculated for each sample by the following method:

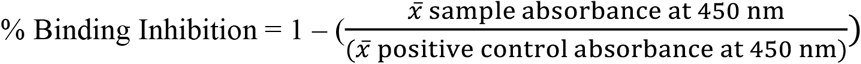

### Data Analysis

GC-FP diagnostic ratio data and percent binding inhibition data for the ACE2 competitive assay were analyzed using GraphPad Prism 8.0 software (ROC analysis, correlation, and statistical analysis).

## RESULTS AND DISCUSSION

### Determination of Human Antibody Response to 2019 SARS CoV2 Vaccination

Dried blood samples from individuals with no known previous COVID-19 infection (n = 52) and individuals two weeks past full vaccination with either Pfizer-BioNTech (n = 17), Moderna (n = 8), or Johnson & Johnson (n = 9) vaccines were tested using the GC-FP assay to determine antibody levels against three SARS CoV2 spike protein antigens (S1, S1S2, RBD) and the nucleocapsid protein (N). As we described in previous work (12), the GC-FP diagnostic ratio provides a quantitative measure of antibody levels and correlates well with established serological techniques such as MIA and ELISA. In the current work, antibody levels in vaccinated individuals were significantly elevated for all spike antigens (Mann-Whitney, p < 0.0001), with highest mean increase in antibody levels observed for the S1 and RBD antigens (supplementary information, Figure S2A). A nominal increase in antibody levels was also observed against the S1S2 antigen and N protein, but reactivity to these antigens was significantly less than for S1 and RBD. Receiver operator characteristic (ROC) analysis on these data (supplementary information, Figure S2B) showed that antibody levels against S1 were diagnostic for vaccination status with 61% sensitivity and 100% specificity, while levels against RBD were diagnostic with 80% sensitivity and 100% specificity. Antibody levels for S1S2 and N resulted in low area under the curve (AUC) results from the ROC analysis and were considered insufficient for diagnostic purposes. Because all three vaccines should only elicit immunological response to the SARS CoV2 spike antigen, the lack of a diagnostic antibody response for the N protein was expected.

Dried blood samples were also tested using GC-FP for ten (10) individuals throughout their vaccination sequence with the Pfizer-BioNTech vaccine (pre-vaccine, at the time of the 2^st^ dose, and 2 weeks after 2^nd^ dose). Antibody levels (as determined by GC-FP detection ratio) were significantly higher for the S1, S1S2, and RBD antigens at both the time of the 2^nd^ dose and 2 weeks after the second dose (Figure 1). As expected, antibodies against the N antigen were not detected at any of these time points, since all three vaccines utilize the spike antigen (and not the N antigen) to induce immune response.

**Figure 1.**
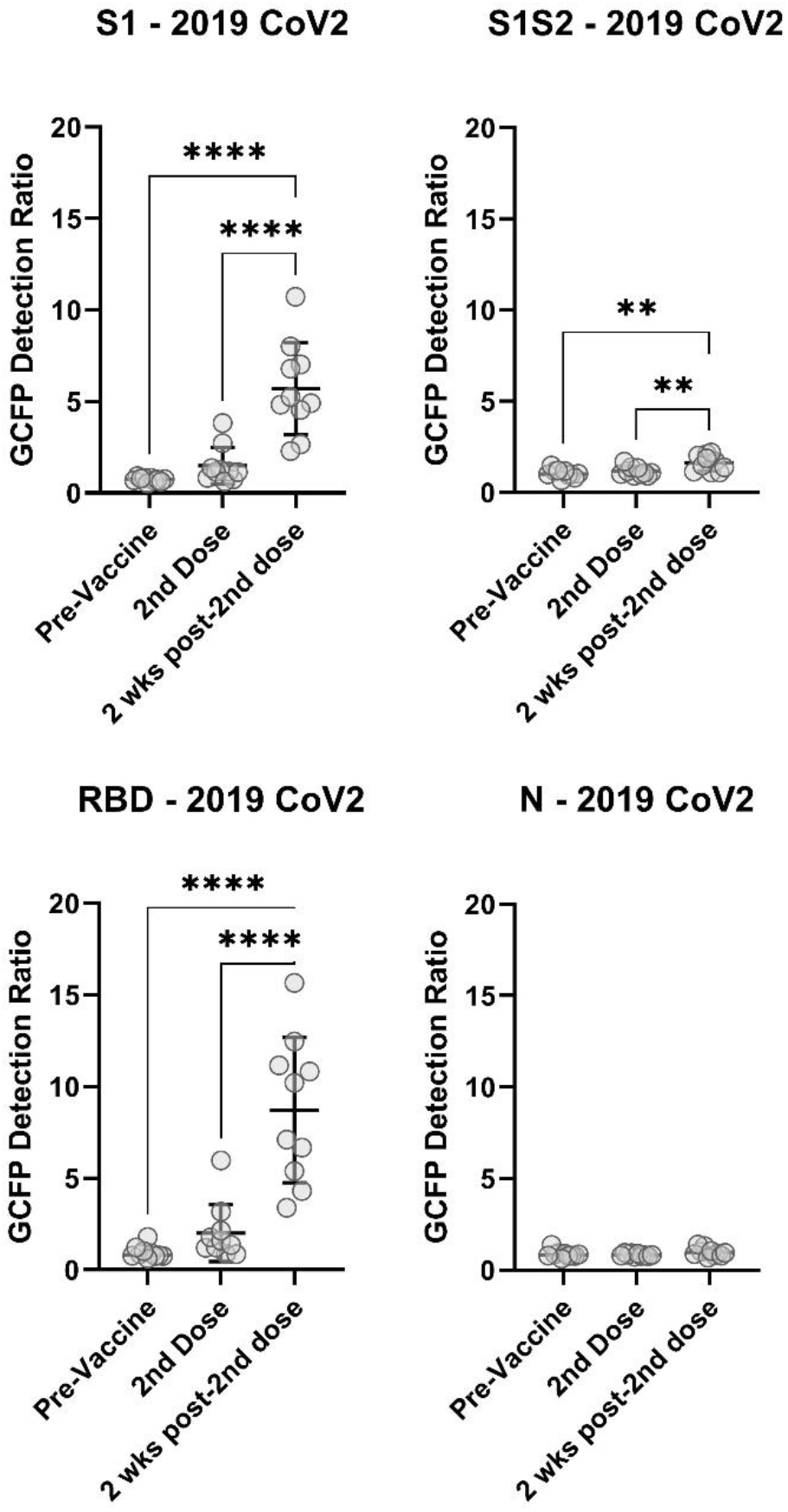
Human IgG levels against SARS CoV2 antigens throughout the vaccination sequence (Pfizer-BioNTech) for10 different subjects, collected pre-vaccination, at the time of the 2^nd^ dose of vaccine, and 2 weeks after the 2^nd^ dose of vaccine. One-way ANOVA followed by Dunnett’s multiple comparison testing was performed (*p = 0.03, ** p = 0.002,*** p = 0.0002, **** p < 0.0001).

GC-FP was also used to detect antibody levels against SARS CoV2 antigens and antigens from variants B.1.1.7 and B.1.351, at additional time points throughout the vaccination sequence (Pfizer-BioNTech) for an individual subject (Figure 2). Increased antibody levels were observed against spike antigens (S1, S1S2, and RBD) within 1 week after the 1^st^ dose. Antibody levels then declined slightly until the 2^nd^ dose, when they increased at both 10 hrs and 1 week post 2^nd^ dose. Finally, levels declined at 2 weeks post 2^nd^ dose. The changes in antibody levels correlate well with what is expected during the vaccination sequence, wherein antibody levels should increase after each dose, but then decline to a stable level over time. Antibody responses to S1 or RBD from CoV2 UK variants were similar as antigens from the original 2019 CoV2, but these responses were dramatically reduced to S. African variants (RBD and S1 from strain B.1.351) throughout the vaccination course. These data illustrate the potential to quantitatively measure antibody response to vaccination with high resolution (through time).

**Figure 2.**
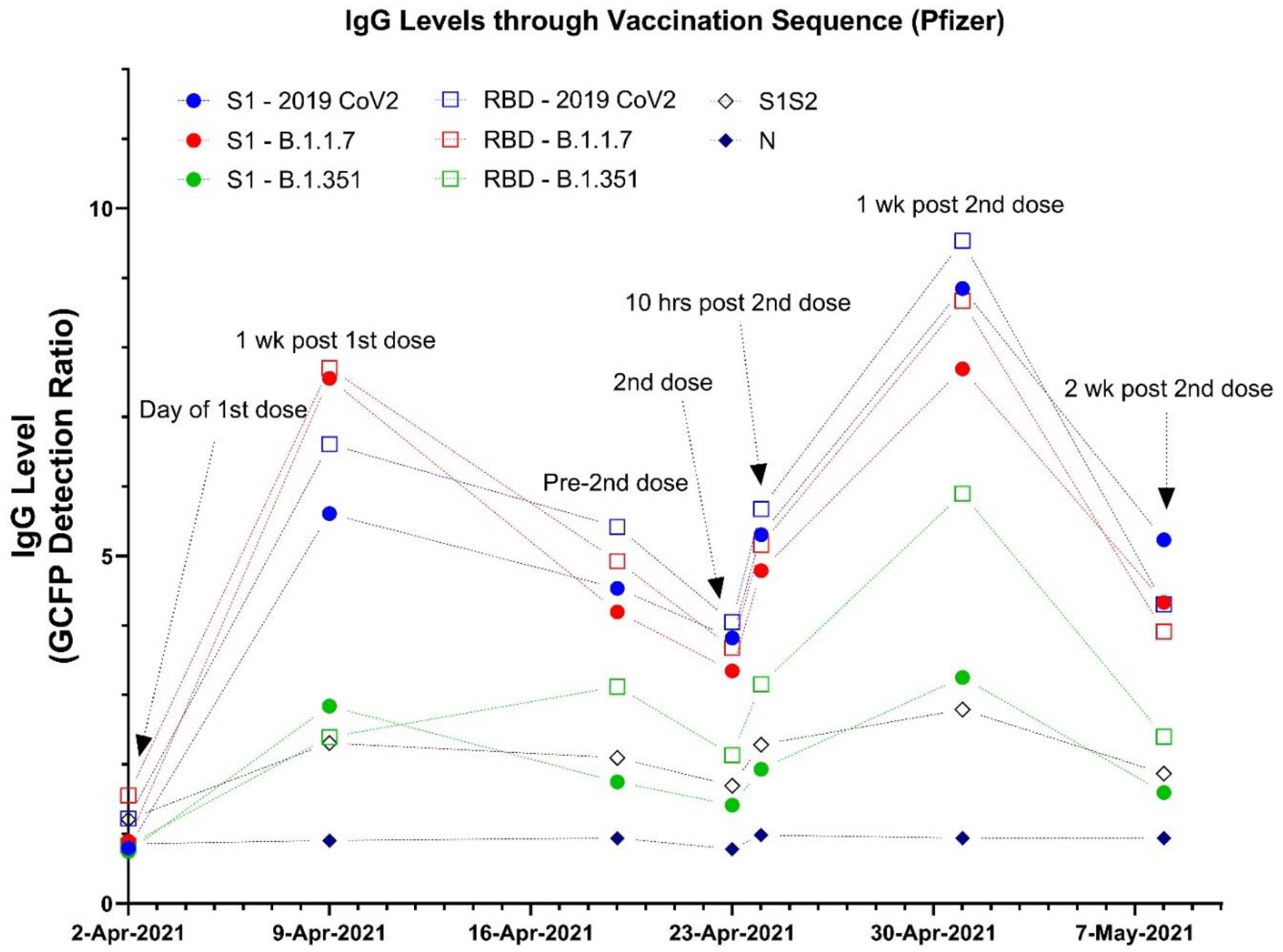
Human IgG levels for a single individual over the course of vaccination with the Pfizer-BioNTech vaccine, measured with GC-FP.

### Determination of Human Antibody Binding to 2019 SARS CoV2 Antigens and its Variants

GC-FP testing was performed on serum from acutely infected (hospitalized) individuals and dried blood samples from individuals who were: 1) <u>Uninfected/pre-vaccine</u> - had no known prior infection with COVID-19 and were not vaccinated, 2) <u>Hospitalized</u> - infected with COVID-19 and hospitalized due to infection, 3) <u>Non-hospitalized</u> - were infected with COVID-19 and at least 4 weeks post recovery, 4) <u>Vaccinated (Pfizer, Moderna, J&</u>J - were at least 2 weeks past final dose of Pfizer-BioNTech, Moderna or Johnson & Johnson vaccine, or 5) <u>CoV2 positive & Vaccinated</u> - previously infected with COVID-19 and then fully vaccinated with Pfizer-BioNTech or Moderna vaccine. Results of this study are shown in Figure 3. As compared to uninfected/unvaccinated individuals, vaccinated individuals (with and without prior infection) had significantly higher antibody levels against S1 and RBD antigens. Acutely infected (hospitalized) individuals had elevated antibody levels for S1S2 and N antigens, while non-hospitalized patients only showed increased antibody levels against N. Notably, individuals who were vaccinated after prior infection had the highest mean antibody levels against S1 and RBD. This is consistent with the fact that these individuals were effectively exposed to the spike antigen at three different time points (during infection and at the 1^st^ and 2^nd^ doses of vaccine).

**Figure 3.**
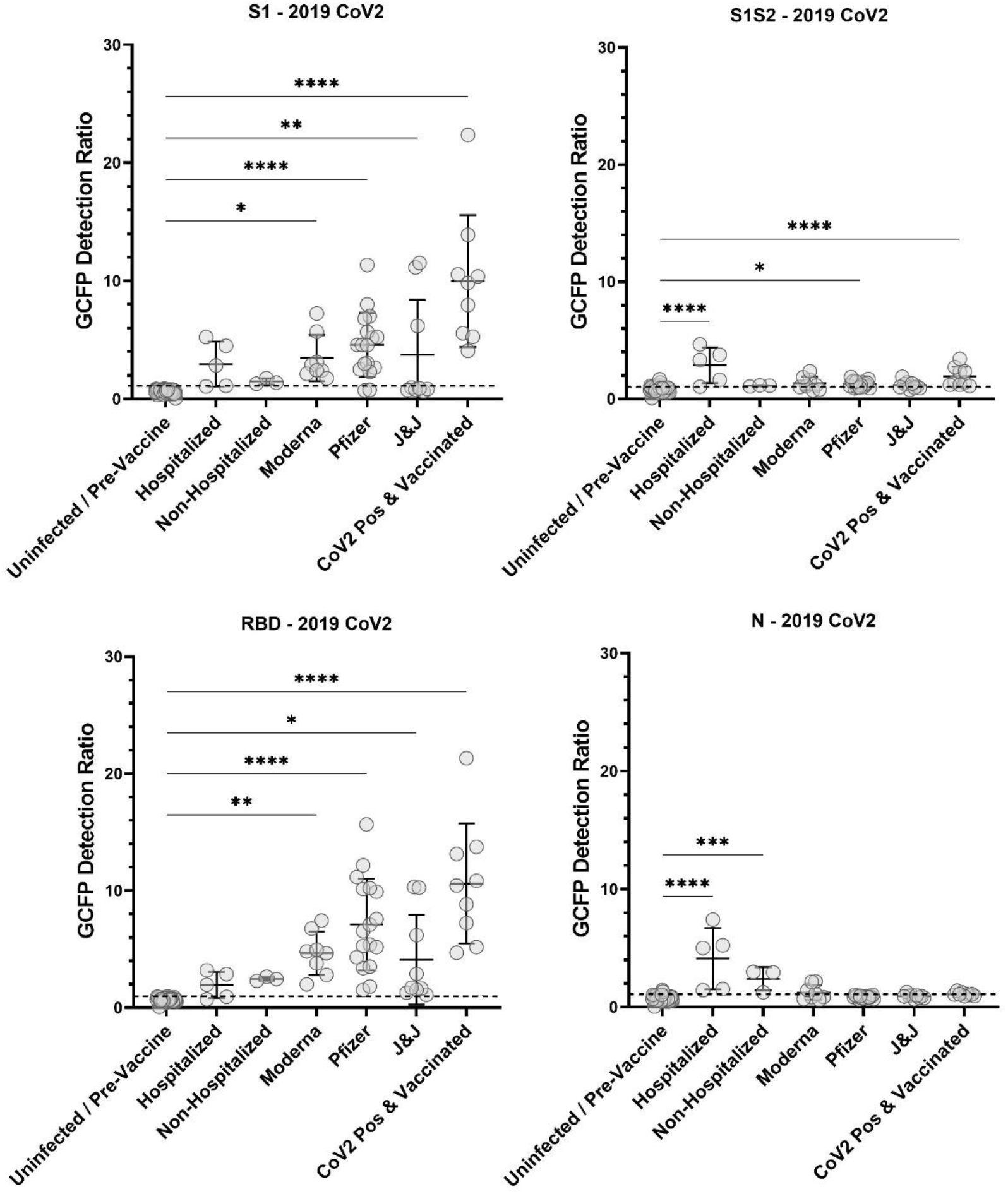
IgG levels against SARS CoV2 antigens for uninfected, previously infected, and vaccinated individuals. Uninfected samples were collected prior to vaccination, from individuals who reported no prior COVID-19 symptoms, and tested negative via PCR and/or antibody testing (n = 42). Other samples were from PCR confirmed COVID-19 positive subjects who were hospitalized (n = 3), PCR confirmed COVID-19 positive subjects who were not hospitalized CoV2 (n = 5), and previously COVID-19 positive subjects who received subsequent vaccination (n = 9). Samples were also collected from subjects who were at least 2 weeks past full vaccination with Pfizer-BioNTech (n = 17), Moderna (n = 8), or 2 weeks after receiving the Johnson & Johnson vaccine (n = 9). One-way ANOVA followed by Dunnett’s multiple comparison testing was performed (*p = 0.03, ** p = 0.002, *** p = 0.0002, **** p < 0.0001).

Between individual subjects, antibody levels were highly variable, regardless of the mode of COVID-19 exposure or vaccination status (Figure 3 and supplementary data Figure S3). To account for these differences, and to elucidate relative antibody binding levels to variant antigens vs. 2019 SARS CoV2 antigens, the fold-difference (binding to variant antigen vs. 2019 SARS CoV2 antigen) in antibody levels between variant antigens and the 2019 SARS CoV2 antigens was plotted (Figure 4). For all vaccinated individuals, antibody binding to B.1.351 antigens (both RBD and S1) was reduced vs. 2019 SARS CoV2 antigens. Antibody binding to B.1.1.7 antigens was either equivalent to, or slightly higher than 2019 SARS CoV2 antigens for these same individuals. For previously infected individuals (hospitalized and non-hospitalized) there were only minor differences in antibody levels for variant antigens vs. 2019 SARS CoV2 antigens (p > 0.03 or not significant).

**Figure 4.**
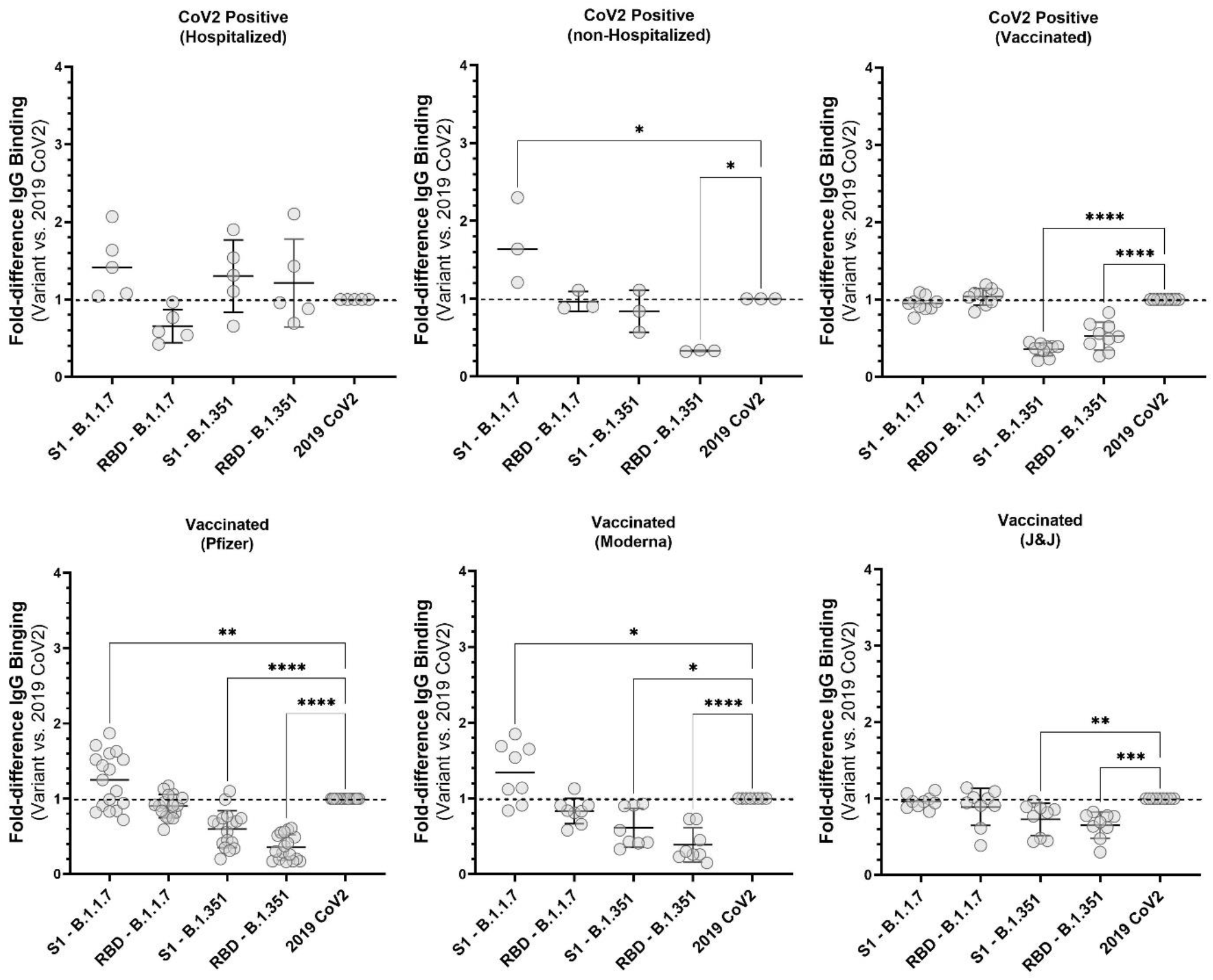
Fold-difference in antibody levels against antigens from SARS CoV2 variant strains B.1.1.7 and B.1.351 vs. antigens from the original 2019 SARS CoV2 strain. Samples included those from individuals who were hospitalized (n = 3); non-hospitalized CoV2 positive (n = 5); previously CoV2 positive with subsequent vaccination (n = 9); and at least 2 weeks past vaccination with Pfizer-BioNTech (n = 17), Moderna (n = 8), or Johnson & Johnson vaccine (n = 9). One-way ANOVA followed by Dunnett’s multiple comparison testing was performed (*p = 0.03, ** p = 0.002, *** p = 0.0002, **** p < 0.0001).

To determine if these results were due to differences in antibody affinity towards variant vs. 2019 SARS CoV2 antigens, we compared the GC-FP results to a competitive ELISA in which blood samples were mixed with ACE2 receptor protein (the cellular target of SARS CoV2) and allowed to competitively bind to 2019 SARS CoV2 RBD antigen, or the B.1.1.7 and B.1.351 RBD variants. Competitive ELISA results were similar to GC-FP results (Figure 5, A&B), showing both significantly reduced antibody binding (GC-FP) and reduced binding inhibition (competitive ELISA) for the B.1.351 RBD variant. When quantitative dilution testing was performed for a single Pfizer-BioNTech-vaccinated individual and a single acutely infected (hospitalized) individual, significant differences in binding inhibition were observed for both B.1.1.7 and B.1.351 RBD variants (Figure 5,C&D). When the fold-difference in antibody binding (GC-FP) and percent binding inhibition (competitive ELISA) were compared, there was close correlation between the two methods (Figure 5, E&F, and supplementary data Figure S4). Values plotted in Figure 5, E&F were shown to be correlated (Pearson r = 0.89, p = 0.02, supplementary Figure S4), suggesting that GC-FP has utility for assessing relative antibody binding levels and/or avidity to antigens from 2019 SARS CoV2 and variant strains of the virus.

**Figure 5.**
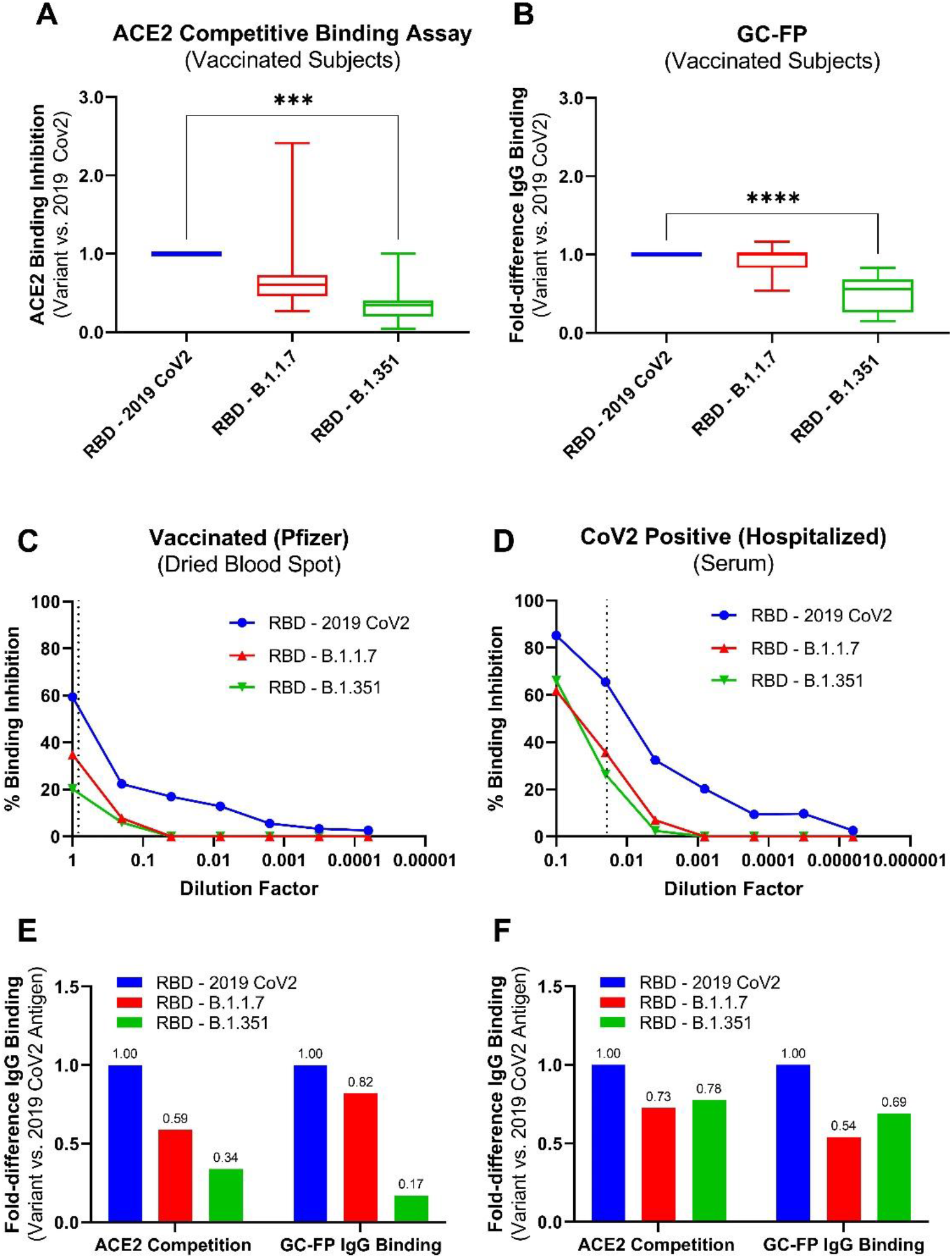
A&B) IgG levels from dried blood spots measured by GC-FP diagnostic ratio compared to competitive ELISA by eluate from the same dried blood spot samples. Both GC-FP and ACE2 competitive binding were performed for RBD antigen from the original 2019 SARS CoV2 and the variant strains B.1.1.7 and B.1.351. Testing was performed with dried blood spots collected from vaccinated subjects (3 Pfizer-BioNTech, 3 Moderna, 2 Johnson & Johnson) and subjects who were both previously infected and then vaccinated with Pfizer-BioNTech or Moderna vaccines (n = 3). One-way ANOVA followed by Dunnett’s multiple comparison testing was performed (*p = 0.03, ** p = 0.002, *** p = 0.0002, **** p < 0.0001). Percent ACE2 binding inhibition from competitive ELISA assay is shown for a dried blood spot sample (C) from a Pfizer-BioNTech vaccinated subject and blood serum from a hospitalized, COVID-positive subject (D). Vertical dotted lines represent the dilution factor used in the corresponding GC-FP test for each sample. Fold difference in binding inhibition and fold difference in GC-FP diagnostic ratio was plotted for variant antigens (RBD B.1.1.7 and RBD B.1.351) vs. RBD 2019 CoV2 (E & F).

## DISCUSSION

We previously demonstrated that GC-FP is a rapid and accurate technique for detection of antibodies that result from COVID-19 infection (12). In the current study, we extended this work to show that GC-FP can measure the level of antibodies resulting from vaccination, and that it can quantitatively measure the increase in antibody levels during the course of vaccination for multiple target antigens. Using ROC analysis, GC-FP diagnostic ratio thresholds could be established to yield a clear cut-off for determining whether or not an individual has been vaccinated. When using the RBD antigen, this results in 80% sensitivity and 100% specificity for all three of the currently approved vaccines (Pfizer-BioNTech, Moderna and Johnson & Johnson). Furthermore, our results show that antibody levels can be measured with high resolution throughout the course of vaccination, making it a useful tool to track the progression of an individual’s serological response to a vaccine. Highlighting the sensitivity of the GC-FP approach, we were able to detect increasing levels of antibodies within just 10 hrs of an individual’s second dose of the Pfizer-BioNTech vaccine (Figure 2).

Beyond determination of vaccination status and serological response to vaccination, the multiplexed nature of GC-FP makes it highly amenable to measuring antibody binding to multiple antigens from both the original 2019 SARS CoV2 virus and the emerging variants of this virus. This is extremely important as variants of the virus continue to emerge (5, 6). For example, GC-FP results indicate reduced antibody binding to the RBD and S1 antigens of the B.1.351 (S. African) variant, which was further confirmed through competitive ELISA-based testing (Figure 5). When individual samples were evaluated by GC-FP and competitive ELISA, reduced antibody binding to antigens from both the B.1.1.7 (UK) and B.1.351 variants was observed.

The results of the present study, together with our previous demonstration of GC-FP for COVID-19 antibody detection (12) compare favorably to similar studies using multiplexed, array-based techniques. Using a plasmonic-based approach, Liu et al. demonstrated high throughput detection of COVID-19 antibodies from human serum and saliva (17). This approach requires significantly more time than GC-FP (∼2 hrs vs. 30 min) but it has the advantage of measuring the relative avidity of antibodies towards target antigens. In another study, Swank et al., demonstrated an elegant, high throughput microfluidic approach to COVID-19 antibody detection. While this approach enables extremely high throughput, it is limited in the number of target antigens that can be implemented, and requires complicated printing of human blood samples to prepare detection chips.

In the work presented here, we have demonstrated that GC-FP based, multiplexed detection of antibody binding from human blood is a useful tool for determining an individual’s response to vaccination and the relative binding of antibodies to variants of the 2019 SARS CoV2 virus. When considering the rapid time to result (30 min) for the GC-FP assay and the ability to target a large number of antigens at once, this represents a powerful tool for continued management of the global COVID-19 pandemic, and with broad applicability to other diseases and vaccines.

## Data Availability

Data is available upon request.

